# Behavioral economic strategies to increase naloxone acquisition and carrying

**DOI:** 10.1101/2024.01.18.23296155

**Authors:** Rachel Feuerstein-Simon, Abby Dolan, Meghana Sharma, Margaret W. Lowenstein, Alison M. Buttenheim, Zachary F. Meisel, Carolyn C. Cannuscio

## Abstract

**Background:** The opioid overdose crisis claimed over 80,000 American lives in 2019, with opioids implicated in the majority of these deaths. The COVID-19 pandemic has exacerbated the crisis, with challenges arising from the increased use of fentanyl, synthetic opioids, and combined opioid-stimulant substances. Urgent strategies are required to mitigate drug-related harms in the context of an unsafe drug supply.

**Objective:** This paper presents the findings of two consecutive randomized controlled trials conducted in Philadelphia, aiming to assess the effectiveness of behavioral economic interventions in promoting naloxone acquisition and carrying among adults.

**Methods:** The trials focused on increasing naloxone acquisition (Study A) and promoting naloxone carrying (Study B) among participants who completed community-based overdose recognition and reversal training. Participants were randomized into three arms: those receiving text message nudges, those signing commitment contracts, and a control group receiving only in-person overdose reversal training. Data collection utilized a web-based platform, and participants were compensated upon study completion.

**Results:** In Study A, participants were encouraged to acquire naloxone, and the primary endpoint was naloxone acquisition within four weeks post-training. Results showed that around one in five participants acquired naloxone, regardless of the intervention arm, indicating limited effectiveness of behavioral strategies in promoting naloxone acquisition.

In Study B, where all participants received naloxone by default, the primary endpoint was the consistency of naloxone carrying. Participants had naloxone on hand at approximately half of the eight unannounced check-ins, with no significant differences between intervention groups and the control group.

**Conclusion:** Naloxone distribution by default emerges as a promising strategy to increase naloxone possession and save lives amidst the opioid overdose crisis. Addressing structural barriers, including cost and pharmacy availability, is essential. Additionally, interventions should consider leveraging motivations such as altruism and regret aversion to encourage naloxone carrying.

## MANUSCRIPT

Over 80,000 Americans died from drug overdose from May 2019 - May 2020, and opioids were implicated in the majority of those deaths.^1^ In addition, the burden of overdose mortality has increased during the COVID-19 crisis, compounding the escalating challenges associated with more common use of fentanyl, other synthetic opioids, and combined opioids and stimulants.^1^ Effective strategies are urgently needed to mitigate the harms of drug use–amidst an unsafe drug supply–and to reduce drug-related mortality.^1,2^

Among interventions to decrease opioid-related mortality, wider availability of naloxone, the opioid overdose antidote, is estimated to hold the most promise, with the potential to save 21,200 lives over the next ten years.^3^ Naloxone, often known by the brand name Narcan (a nasal spray formulation), is easy to use, highly effective, and has few serious side effects beyond opioid withdrawal symptoms.^4^ Multiple studies have documented that first responders, including EMS and police, as well as laypeople, can be trained to recognize and reverse opioid overdoses, thereby saving lives.^5-9^

Although bystanders were present at half of fatal opioid overdoses in 2016, they attempted to reverse those overdoses using naloxone in fewer than 5% of cases.^10^ These findings suggest a large unmet need to improve naloxone availability and use at the time of overdose. Early efforts to increase naloxone access focused on distribution to people who use drugs (PWUD), as well as police, EMS, and other first responders, with subsequent mortality reductions.^6^ In addition, a major effort to distribute naloxone to PWUD and their social networks resulted in marked reductions in opioid overdose fatalities in Massachusetts communities.^5^ We must build on this success and close the remaining major gap in naloxone uptake and utilization.^11^ For example, only a small fraction (1.5%) of insured patients at high risk for overdose were prescribed naloxone as recently as 2016.^12^ Even with the recent shift to over-the-counter status for naloxone, there are open questions regarding challenges in cost, access, stigma, and consistency of carrying–as well as challenges to people who use drugs alone.

In order to close the gap in naloxone use and save lives, strategies must be identified to increase carrying and use of naloxone during overdose events. Behavioral economic approaches have been employed successfully by policymakers, researchers, and public health practitioners to increase adoption of healthy behaviors.^13-20^ Behavioral nudges--strategies designed to encourage desired behaviors--have been shown to encourage smoking cessation, weight loss, and adherence to physician recommendations after emergency room visits.^15,16,18^ Commitment contracts, which are pledges designed to activate personal and social accountability, have helped promote smoking cessation.^21^

Informed by the success of behavioral nudges in other contexts, the authors conducted two sequential randomized controlled trials to test the effectiveness of text message reminders (Study A) and commitment contracts (Study B) in promoting naloxone acquisition and naloxone carrying among adults in Philadelphia.

## Methods

### Rapid cycle trials to increase naloxone acquisition (Study A) and carrying (Study B)

We conducted two randomized controlled trials to test behavioral economic strategies to increase naloxone acquisition (Study A) and to promote naloxone carrying (Study B). The studies were conducted between February 2019 and August 2019. Several features were shared across both studies, such as the inclusion criteria (see Table A for key study features). Both studies enrolled adults ages 18 and older who lived or worked in the Philadelphia area, participated in community-based overdose recognition and reversal training, and could understand and complete the training and associated study instruments in English. For both studies, participants were recruited using a range of strategies, including outreach through community listservs, social media, word-of-mouth, and public poster announcements in Philadelphia. Sample sizes were determined by funds available rather than by *a priori* hypotheses and power calculations. Participants were compensated $20.00 upon completion of the study.

Study staff obtained written consent from participants over the phone using Way to Health, a web-based platform designed to facilitate studies and engage research participants.^22^ All participants completed a baseline survey assessing 1) prior experience with witnessing overdose, 2) motivations for training participation, 3) readiness for opioid overdose reversal (including self-efficacy, knowledge, and intentions), and 4) stigma regarding people who use opioids. All surveys were deployed electronically using the Way to Health platform.

In both studies, all participants completed a 1-hour in-person overdose recognition and reversal course, which addressed the following topics: the scope and nature of the opioid overdose epidemic nationally and locally in Philadelphia, signs of opioid overdose, strategies for approaching or interacting with a person who is suspected of having overdosed, administration of naloxone in its nasal spray formulation, what to expect after administering naloxone, legal considerations and Good Samaritan protection, and how and where to acquire naloxone.

Both studies were three-arm trials in which participants were randomized in equal proportions to each arm using Way to Health. In each study, participants in Arm 1 received an intervention that included 14 text message “nudges” tailored to address specific barriers and concerns regarding naloxone acquisition and carrying. These barriers were identified using qualitative interviews conducted by the authors in prior studies.^23^ Examples included optimism bias (e.g., beliefs that participants were unlikely to encounter someone who had overdosed in their daily lives) and identity bias (e.g., beliefs that participants were not the type of people who could save a life). Participants in Arm 2 did not receive text message prompts but instead were asked to sign commitment contracts, further described for each study in the sections below. In each study, participants in Arm 3 were the usual care control group, and they received only the in-person overdose reversal training, without text message nudges or commitment contracts. Participants in both studies were followed for four weeks post-training using the Way to Health platform, which automated text message correspondence and data collection.

### Study A: Behavioral strategies to increase naloxone acquisition

For Study A, since the primary endpoint was naloxone acquisition, only people who had not previously acquired naloxone were eligible to enroll in the trial. All training participants were encouraged to acquire naloxone, either through pharmacies or through local harm reduction organizations. During the training, the study team offered guidance regarding Pennsylvania naloxone policies, locations of pharmacies that typically stocked naloxone, and the name and contact information for a harm reduction organization that provides naloxone at a reduced price.

The primary endpoint was assessed at eight unannounced check-ins during the four weeks following the training. Participants in all three treatment arms were sent text messages asking, “Have you acquired naloxone? If no, text “NO” to this number. If yes, please respond within 2 hours with a photo of your naloxone and [a specific code word of the day] written on a piece of paper visible in the photo.” This photo-documentation method was used in prior behavioral economic trials of epinephrine carrying and was shown to be a low-burden measurement approach.^24^ The two-hour window was intentionally broad to allow participants to respond without interruption of activities, such as driving. The primary endpoint was achieved if the participant succeeded in documenting their possession of naloxone at any one of the eight check-ins.

### Study B: Behavioral strategies to encourage naloxone carrying

In Study B, all overdose reversal training participants were given naloxone by default. The primary endpoint was consistency of naloxone carrying during the four-week period following the training. The endpoint was ascertained using photo-documentation, as in Study A. Responses were considered successful if they were received by the study team within two hours of the text message check-in.

### Analysis

All analyses were conducted using R version v.1.1.45. Descriptive statistics were calculated to characterize the study population, with chi-square tests used to compare proportions and t-tests used to describe differences in means across treatment arms within each study. To analyze the primary endpoint for Study A, naloxone acquisition in the four weeks following training (yes/no), we compared proportions across study arms using chi-square tests.

In Study B, we measured the consistency of naloxone carrying as the proportion of eight unannounced text message check-ins at which the participant successfully documented that they were carrying naloxone by replying with a photograph that included the naloxone and a code word of the day. We conducted chi-square tests to compare differences in proportions of successful check-ins across treatment arms.

Exploratory analyses were conducted to compare results across Study A and Study B. This post-hoc comparison was conducted to assess differences in naloxone carrying among groups that had or had not been given naloxone by default during the training. Specifically, we compared the proportion of participants in each study who were in possession of naloxone for at least one check-in during the four-week follow-up period. Participants in Study B were all given naloxone by the study team upon completion of the overdose reversal training.

Additionally, we conducted post-intervention surveys, which included further questions about participant perceptions of the interventions (text messages and commitment contracts), barriers to success in acquiring or carrying naloxone, and feedback regarding the overdose reversal training. All follow-up surveys were sent within five weeks of training completion.

The study was reviewed and approved by the University of Pennsylvania Institutional Review Board and registered on www.clinicaltrials.gov (ID: NCT06064981).

## Results

### Study A, Naloxone Acquisition

Baseline characteristics of Study A participants (n=53) are shown in Table 2. In Study A, training participants were encouraged to acquire naloxone (primary endpoint), either at a local pharmacy or through a public health or harm reduction organization, within the four weeks following the training. None of the participants had acquired naloxone prior to the study.

**Table 1.** Key study features across Study A and Study B.

| ***Table 1. Key study features across Study A and Study B*** |  |  |
| --- | --- | --- |
| ***Studies*** | **Study A: Testing behavioral economic strategies to increase naloxone acquisition.** | **Study B: Testing behavioral economic strategies to promote naloxone carrying.** |
| ***Recruitment*** | Participants were recruited using a range of strategies, including outreach through community listservs, social media, word-of-mouth, and public poster announcements in Philadelphia. |  |
| ***Sample*** | Studies included participants ages 18 and older who lived or worked in the Philadelphia area, participated in a community-based overdose recognition and reversal training, and could understand and complete the training and associated study instruments in English. |  |
| ***Randomization*** | Participants were randomized into one of three arms using the Way to Health platform. |  |
| ***Study Arms & Intervention*** | All study participants completed a 1-hour in-person overdose recognition and reversal course. Following the course, participants received one of the following interventions:<br>Arm 1. Over the 4 weeks following the training, participants received 14 text message “nudges,” tailored to address specific barriers and concerns regarding naloxone acquisition and carrying.<br>Arm 2. Participants were asked to sign contracts indicating a commitment to: acquiring naloxone (Study A) or consistently carrying naloxone (Study B).<br>Arm 3. Participants received usual care (i.e., in-person overdose reversal training only, without nudges or commitment contracts). |  |
| ***Primary endpoint and analysis*** | **Naloxone acquisition**, measured as the successful completion of at least one text message check-in at which the participant successfully documented that they had acquired naloxone, by replying with a photograph that included the naloxone and a code word of the day. | **Consistency of naloxone carrying**, measured as the proportion of eight unannounced text message check-ins at which the participant successfully documented that they were carrying naloxone, by replying with a photograph that included the naloxone and a code word of the day. |
| ***Exploratory endpoints*** | Proportion of participants in each study who were successful on at least one check-in during the four-week follow-up period after the overdose reversal training, indicating that they were in possession of naloxone. |  |
|  | Barriers to **acquiring** naloxone.<br>Perceived importance of **acquiring** naloxone.<br>Perceived prevalence of naloxone **acquisition**. | Barriers to **carrying** naloxone.<br>Perceived importance of **carrying** naloxone.<br>Perceived prevalence of naloxone **carrying**. |
| ***Study platform*** | Randomization, assessments, surveys, and text messages were deployed using the Way to Health automated data collection, communication, and analysis platform. |  |

**Table 2.** Study A (objective: naloxone acquisition) participant characteristics (n=53)

|  | Text Message<br>Nudges<br>n=17 | Commitment<br>Contract<br>n=20 | Control<br>n=16 |
| --- | --- | --- | --- |
| **Reason for attending** |  |  |  |
| General interest | 76% | 95.0% | 93.8% |
| Frequent encounters with people at risk for overdose | 59% | 30.0% | 43.8% |
| Need for training related to employment role | 18% | 45.0% | 25.0% |
| Friend/family at risk | 29% | 10.0% | 6.3% |
| Personal risk | 0% | 5.0% | 0.0% |
| **Witnessed an overdose prior to training** | 18% | 30.0% | 37.5% |
| **Reversed an overdose prior to training** | 0% | 5.0% | 0.0% |
| **Race/Ethnicity** |  |  |  |
| White/Caucasian | 94.1% | 55.0% | 81.3% |
| Asian | 0.0% | 15.0% | 18.8% |
| Black | 0.0% | 15.0% | 0.0% |
| Latino | 5.9% | 0.0% | 0.0% |
| More than one race | 0.0% | 15.0% | 0.0% |
| **Gender** |  |  |  |
| Female | 76% | 55.0% | 56.3% |
| Male | 18% | 35.0% | 43.8% |
| Other | 6% | 5.0% | 6.3% |
| **Age (mean)** | 29 | 29 | 30 |

While none of the study participants owned naloxone at baseline, across all three study arms, approximately one in five participants had acquired naloxone by the end of the study period. Naloxone acquisition rates are reported in Table 3. In the control arm, whose members completed the training but otherwise did not receive additional behavioral economic intervention, 19% had acquired naloxone by week four post-training. Acquisition rates were not significantly different in the commitment contract arm (20%) and in the text message nudge arm (24%). These results suggest that, while about 1 in 5 training participants did take the steps necessary to acquire naloxone, the added behavioral economic strategies were not more effective than training alone in encouraging naloxone acquisition.

**Table 3.** Study A: Proportion of participants who acquired naloxone within four weeks of training*†.

| Arm | Acquired naloxone |
| --- | --- |
| Message intervention | 24% |
| Commitment contract | 20% |
| Control | 19% |

\*No statistically significant differences between each intervention and the control arm.
† At baseline no Study A participant owned naloxone, consistent with the inclusion criteria for this study.

In open-ended questions, participants were asked about challenges related to acquiring naloxone. Overwhelmingly, the main barrier to acquisition was cost, which was mentioned by more than half (54%) of participants who had not acquired naloxone. 96% of participants who responded to the post-survey said it was either “Important” or “Very Important” to carry naloxone. 76% said it was “Uncommon” or “Very Uncommon” for people in Philadelphia who are not first responders to carry naloxone. Participants told an average of five additional people about their experience at the training.

### Study B, Naloxone Carrying

In Study B, all training participants were given naloxone, by default, and the primary endpoint was consistency of carrying naloxone in the four week period following the training. Baseline characteristics of Study B participants (n=84) are summarized in Table 4.

**Table 4.** Study B (naloxone carrying) participant characteristics (n=84)

|  | Test<br>Message<br>Nudges<br>n=28 | Commitment Contract<br>n=27 | Control<br>n=29 |
| --- | --- | --- | --- |
| **Reason for attending** |  |  |  |
| General interest | 93% | 93% | 90% |
| Need for training related to employment role | 25% | 19% | 21% |
| Friend/family at risk | 18% | 15% | 3% |
| Personal risk | 0% | 0% | 0% |
| **Witnessed an overdose prior to training** | 21% | 7% | 14% |
| **Reversed an overdose prior to training** | 7% | 0% | 3% |
| **Race/Ethnicity** |  |  |  |
| Asian | 29% | 22% | 24% |
| Black | 11% | 7% | 10% |
| Latino | 7% | 7% | 0% |
| White/Caucasian | 43% | 56% | 59% |
| **Gender** |  |  |  |
| Female | 79% | 67% | 76% |
| Male | 18% | 33% | 21% |
| Other | 4% | 0% | 3% |
| **Age (mean)** | 29 | 31 | 31 |

Results of Study B are summarized in Table 5, showing that, across all three study arms, participants had their naloxone on hand at approximately half of the 8 unannounced check-ins. Text message nudges or commitment contracts were not significantly associated with better performance on the primary outcome, consistency of naloxone carrying.

**Table 5.**
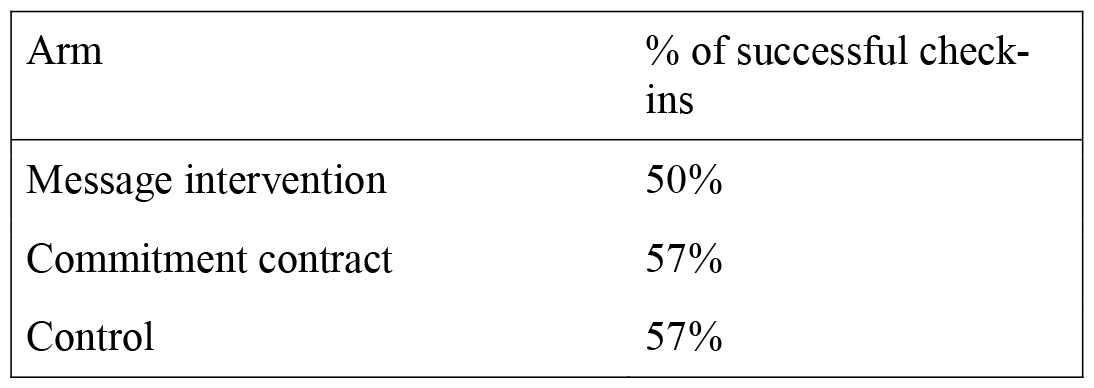
Proportion of 8 check-ins at which participant successfully demonstrated that they were in possession of naloxone, measured over a four-week period following training.

| Arm | % of successful check-ins |
| --- | --- |
| Message intervention | 50% |
| Commitment contract | 57% |
| Control | 57% |

At the end of the four-week follow-up period, we asked a series of open-ended questions regarding what, if anything, made it challenging for participants to carry the naloxone they had been given during the training. There was no difference in the consistency of naloxone carrying between men and women in this study. However, the hassle associated with where to put the naloxone was cited by 65% of male respondents, who noted that carrying in a pocket is cumbersome, while carrying it in a purse or backpack might be more convenient. Women also noted that they were less likely to have their naloxone when they weren’t carrying a bag or purse.

### Distributing naloxone by default at the training was associated with substantially higher proportion of participants carrying naloxone

We conducted exploratory analyses across Study A and Study B, comparing the proportion of participants in each study who had naloxone on hand at any point during the four-week follow-up period. In this exploratory analysis, demonstrating naloxone possession at any 1 of 8 check-ins was considered a success. We observed marked differences in naloxone possession in Study A, in which participants were encouraged to acquire naloxone on their own, compared to Study B, in which participants were by default given naloxone at the training. Figure 1 summarizes the results of this exploratory comparison.

**Figure 1.**
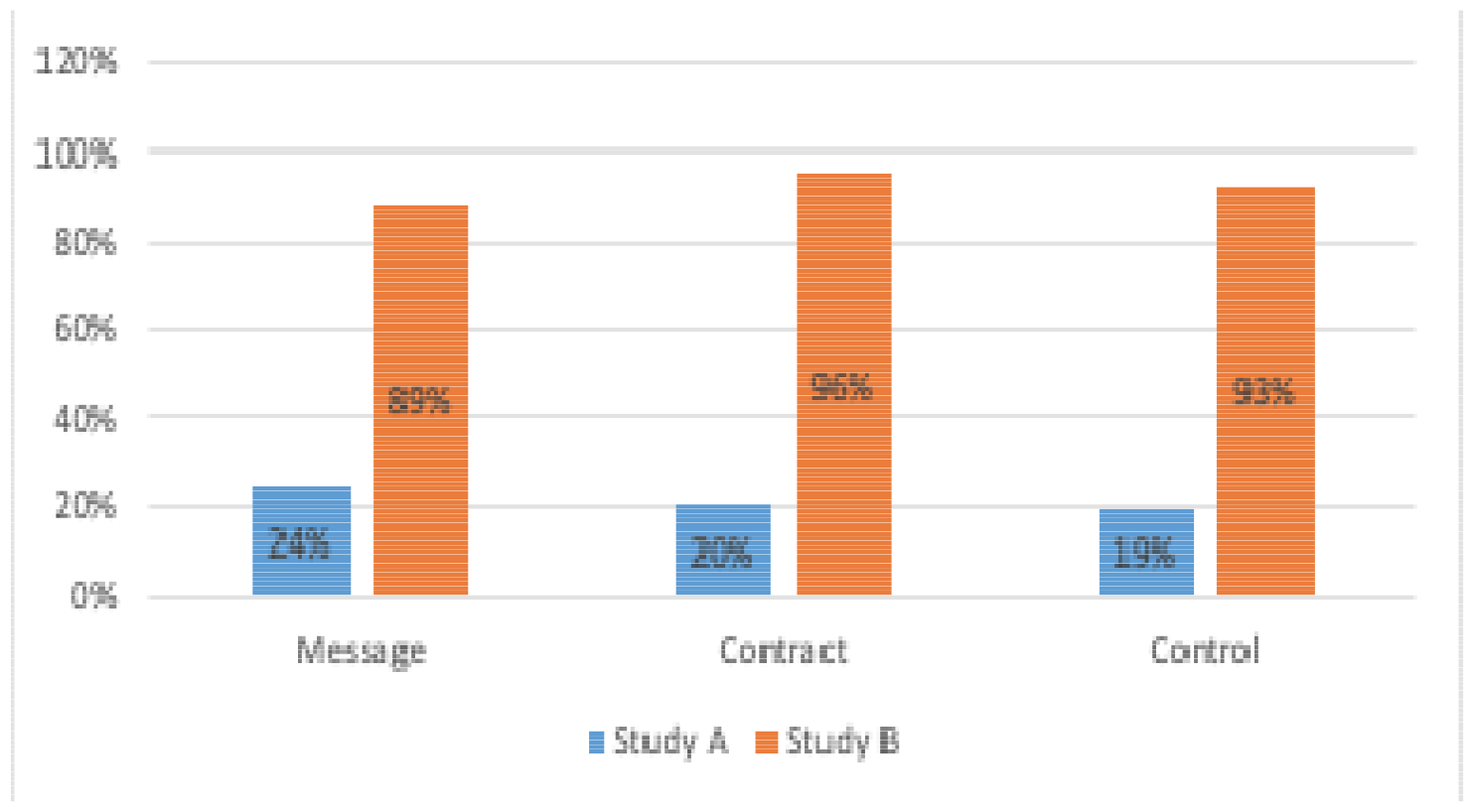
Proportion of study participants who had naloxone in hand in at least one surprise check-in.

Dramatic differences in naloxone possession were observed when we compared participants in Study A, who had to acquire their own naloxone, to those in Study B, who were given naloxone by default at the training.

When participants were expected to acquire their own naloxone, approximately 20% of them successfully acquired the drug and demonstrated that they were carrying it in at least one of 8 check-ins over 4 weeks following training. In contrast, approximately 90% of participants in the naloxone-by-default condition (Study B) demonstrated that they were carrying the medication in at least one of 8 check-ins.

## Discussion

The Surgeon General has called for more Americans to carry naloxone as one step that anyone can take to save lives and limit the toll of the opioid overdose crisis.^25^ This paper offers insights, motivated by behavioral economic principles, that can further inform the opioid overdose crisis response. We advance the public health literature in several key ways.

First, about one in five training participants from Study A acquired naloxone post-training, regardless of their treatment assignment. However, far more participants had naloxone on-hand during follow-up if we distributed naloxone by default at the training. The effect of distributing naloxone (for free) to all training participants far eclipsed the effects of behavioral strategies like text message reminders or commitment contracts, which showed no benefit compared to training alone. Participants were almost four times as likely to successfully document having naloxone at hand if they were given the medication than if they were encouraged to acquire it themselves.

Second, the financial barriers to naloxone acquisition are prohibitive. At the time of this study, naloxone was a prescription medication. Co-pays for naloxone varied by insurance plan, and it could be difficult and time-consuming for people to navigate their insurance company bureaucracies to secure accurate information about costs. For people who were uninsured, the outright cost of naloxone was high, at $140 for two doses of Narcan in most pharmacies.^26-28^ It is also important to note that many people will need to administer and then replace their naloxone, or they may need to deliver multiple doses to reverse a single overdose. With the increasing presence of fentanyl and other synthetic opioids in the drug supply, one 4 mg dose of naloxone is often not sufficient.^29^ Costs will be particularly onerous for people who use drugs, who are likely to assist their peers, or for their close friends and family. This will continue to be true now that naloxone is available as an over-the-counter medication. For people who seek to serve as active bystanders, the cost of naloxone may pose a barrier sufficient to override their desire to help.

Next, cognitive biases and social norms present additional barriers to naloxone acquisition and carrying. Though the text message nudges and commitment contracts were designed to address cognitive biases, they did not achieve further gains in naloxone acquisition or carrying, beyond those observed among participants in the training-only control groups. In retrospect, the training itself addressed both social norms and cognitive biases (e.g., optimism bias that they are unlikely to encounter an overdose) by providing specific data regarding the frequency and geographic distribution of overdose in Philadelphia.

Therefore, a limitation of this study is that there was “crossover,” such that both intervention and control group members were exposed to information that countered cognitive biases. Alternatively, it is possible that the text message “dose” or content were insufficient to produce further gains. Additionally, all participants received text message prompts to document their naloxone, which served as an additional nudge received by both intervention and control group members. Nonetheless, even in Study A, 1 in 5 participants had acquired naloxone post-training, which on a larger scale would translate into appreciably more laypeople equipped to respond to overdoses. Still, this intervention leaves much room for further gains in naloxone uptake, and further interventions should address structural barriers to naloxone access and uptake, including cost and widespread availability at pharmacies, which have been found too often not to stock naloxone–especially in low-income and rural locations.^31^

Finally, as a point of comparison, we can compare results from this study regarding naloxone carrying (in Study B) to results from our prior studies of epinephrine autoinjector carrying among people diagnosed with potentially life-threatening food allergies. In those studies, people who were *themselves* at risk of anaphylaxis and death carried their lifesaving epinephrine auto-injectors only a quarter of the time.^24^ In contrast, people in the current study were far more consistent in carrying naloxone, which they had on hand approximately half the time, once given the drug for free. Notably, naloxone is typically carried to protect the life of *someone else*, since this medication cannot be self-administered during overdose. In this study only one participant noted they were joining the training because they themselves were at-risk for overdose. Therefore, results from this study suggest that training participants may have been motivated by altruism to care for people at risk for overdose. Future interventions to address the opioid crisis could harness altruism as a motivator. An alternative explanation is that participants were motivated by regret aversion–the desire to avoid remorse for action not taken to save a life.32

Naloxone distribution, as a default, should be explored and pursued aggressively in response to the crisis of opioid overdose, which is claiming over 200 lives in the U.S. each day on average. Implementation of naloxone-by-default can be considered in a range of settings and populations. Examples include distribution of naloxone to people in supervised injection facilities and encampments, patients in addiction treatment programs, people visiting the emergency department because of overdose or complications of substance use, law enforcement officers, emergency medical services, firefighters, school and university staff, as well as locations with high rates of drug overdose, such as public libraries, as well as transit hubs.^33^ All avenues should be explored to remove friction from the naloxone acquisition process and increase the availability of naloxone to the people and places that need it, in order to save lives.

## Data Availability

All data produced in the present study are available upon reasonable request to the authors

